# Fading of brain network fingerprint in Parkinson’s disease predicts motor clinical impairment

**DOI:** 10.1101/2022.02.03.22270343

**Authors:** E Troisi Lopez, R Minino, M Liparoti, A Polverino, A Romano, R De Micco, F Lucidi, A Tessitore, E Amico, G Sorrentino, V Jirsa, P Sorrentino

## Abstract

The clinical connectome fingerprint (CCF) was recently introduced as a way to assess brain dynamics and used to predict the cognitive decline in preclinical Alzheimer’s disease. In this paper we explore the performance of CCF in 47 Parkinson’s disease (PD) patients and 47 healthy controls, under the hypothesis that patients would show reduced identifiability as compared to controls, and that such reduction could be used to predict motor impairment. Using source-reconstructed magnetoencephalography signals, we built functional connectomes and observed reduced identifiability in patients compared to healthy individuals in the beta band. Furthermore, we found that the reduction in identifiability was proportional to the motor impairment, assessed through the Unified Parkinson’s Disease Rating Scale, and, interestingly, able to predict it (at the subject level). Along with previous evidence, this paper shows that CCF captures disrupted dynamics in neurodegenerative diseases and is particularly effective in predicting motor clinical impairment in PD.

## INTRODUCTION

Parkinson’s disease (PD), the second most common neurodegenerative disease (after Alzheimer’s disease)^1^, is clinically characterized by the presence of a broad spectrum of both motor and non-motor symptoms (and signs), including neuropsychiatric disturbances, autonomic dysfunctions, and cognitive decline. However, motor impairment remains prominent in the clinical picture^2^. The variability of symptoms across patients, and the presence of a wide spectrum of non-motor symptoms in each patient, suggest that the pathophysiological mechanisms affecting the brain are not restricted to a limited area but, rather, spread well beyond^3^. In fact, predicting clinical impairment has proven elusive so far, perhaps since the correct unfolding of the interactions among brain areas has to be taken into account. Consequently, there is a wide interest in identifying signs of sub-optimal large-scale organization of the brain activity in order to improve diagnosis and clinical management.

Despite brain activity alteration being a robust finding in PD, the description of the alteration regarding the large-scale brain activity is yet to be unanimous. Recently, it was shown that large-scale brain dynamics becomes stereotyped in PD patients, lacking flexibility proportionally to hypersynchronization^4^, which, in turn, is a recurring finding in PD^5–7^. However, several studies also reported a reduction in connectivity related to several different brain areas, including both cortical and subcortical structures^8–12^. In summary, altered brain activity is a main finding in PD, and it is often related to the beta band. We expect that such an alteration may compromise the identification of the individuals based on their connectomes, and that this characteristic may be proportional to the clinical condition of the specific patient. Based on this reasoning, the clinical connectome fingerprint (CCF) has been recently developed to exploit reduced identifiability in patients affected by amnestic Mild Cognitive Impairment (MCI), which is considered the preclinical form of Alzheimer’s disease. In particular, CCF was able to predict the individual cognitive impairment, assessed through Mini-Mental State Examination^13^.

In this work, we hypothesized that the CCF would be a sensible candidate to build a simple marker to predict clinical impairment in PD. To test this hypothesis, we used source-reconstructed magnetoencephalography (MEG) signals. We performed two separate MEG recordings for each subject of both healthy and PD groups. After filtering the source-reconstructed data in the canonical frequency bands, we used the phase linearity measurement (PLM)^14^ to robustly estimate the synchronization between regions. The PLM is a phase-based metric of synchronization insensitive to field-spread, which was shown to be optimal for CCF^13^. Hence, we built frequency-specific adjacency matrices, where rows and columns are brain regions, and the PLM values represent synchronization between them. Then, we estimated the identifiability rate of each group (based on the Pearson’s correlations between the adjacency matrices), for each frequency band. As PD commonly exhibits altered synchronization in the beta band, this was the frequency where we mainly expected reduced identifiability. Furthermore, we compared the similarity of the patients’ connectome (how much each patient is similar to him/herself), with the similarity of the healthy group’s connectomes, thereby obtaining a “clinical fingerprinting’’ score (I*clinical*) for each patient. In other words, we measured how much each patient’s connectome was similar to the average connectome of the controls. Since more stereotyped brain dynamics has been linked to the clinical impairment in PD, we then used the I*clinical* scores to predict motor clinical impairment, as assessed using the Unified Parkinson’s Disease Rating Scale part III (UPDRS). To this end, we built a multilinear regression model to compare the predicted and the observed UPDRS scores^15^.

## RESULTS

We analyzed the fingerprinting of functional connectomes (FCs) (Fig. 1A) in a cohort of 94 subjects, which included 47 healthy subjects (HS) and 47 patients with PD. To this end we used the FCs of each group to build an Identifiability matrix (IM) like in Sorrentino et al.^13^ (Fig. 1B). The FCs were generated using the PLM as a synchrony measure between brain regions, as defined according to the Automated Anatomical Labeling (AAL) atlas, within the five canonical frequency bands. Specifically, two FCs were built for each subject (named *test* and *retest*, respectively) using two acquisitions that were carried out with a 2-minutes break from each other. For each group, the IM was obtained by correlating the FCs *test* with the FCs *retest* of the subjects. Each IM contains information about the similarity between the FCs of the same subject (I-self score, main diagonal elements), and between FCs of different subjects (I-others, off diagonal elements). Furthermore, it was possible to obtain a differential identifiability score (I-diff) computing the difference between I-self and I-others values. Additionally, clinically relevant features have been further investigated using the I*clinical* score of each patient, obtained by building hybrid IM which included mixed data from both healthy individuals and patients (see methods for further details). The I*clinical* score is a representation of the similarity of a patient FC with the healthy individuals FCs.

**Figure 1.**
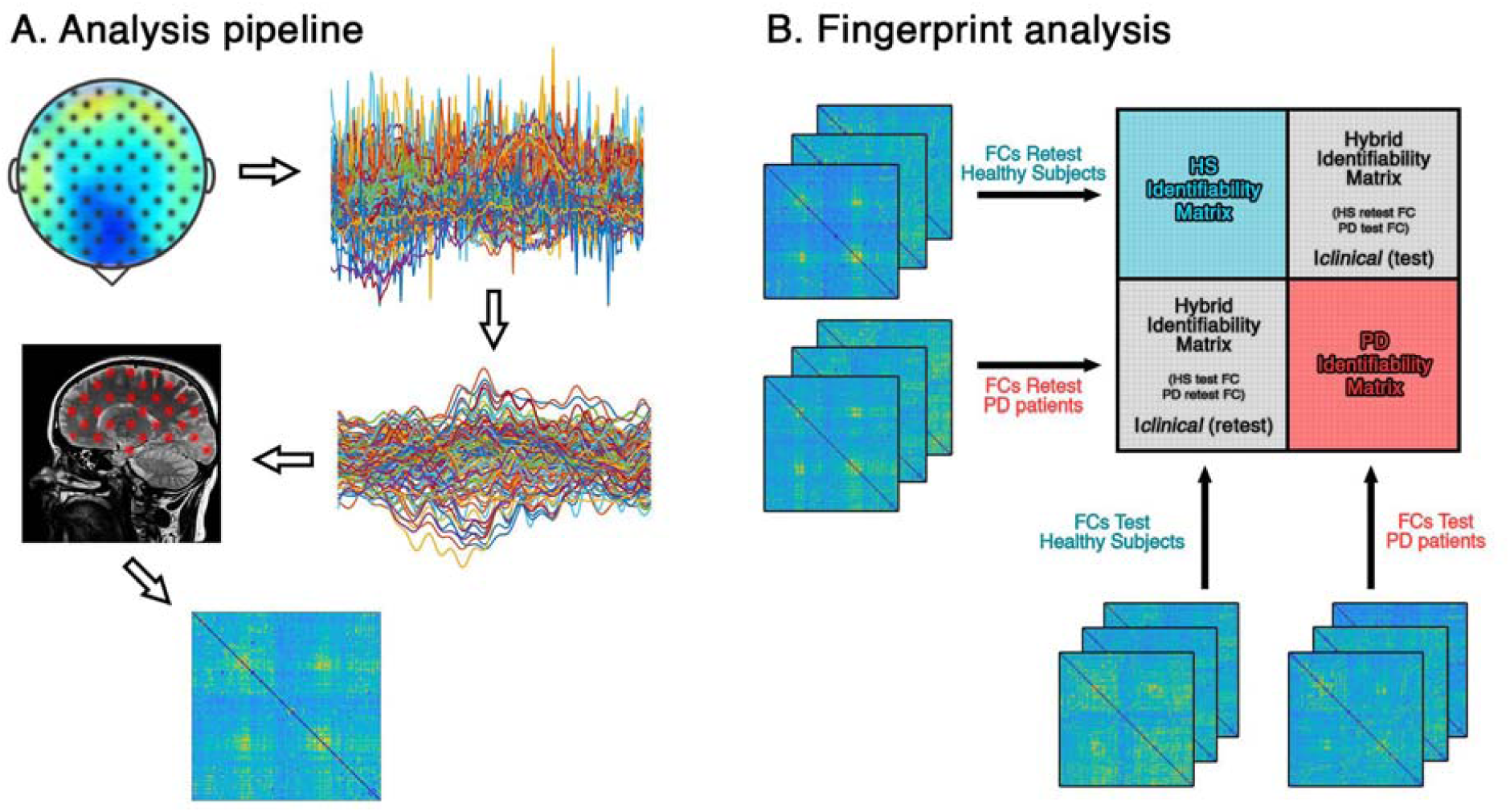
Processing of the functional connectomes and their application for fingerprint analysis. (A) Visual representation of the data analysis pipeline. Through a magnetoencephalography (MEG) system composed of 154 sensors we recorded the magnetic field emitted by neural activity. Noisy MEG signals were cleaned, and artifacts were removed. Source reconstruction (beamforming) was achieved according to the Automated Anatomical Labeling atlas. Connectivity estimation was performed through Phase Linearity Measurement (PLM) algorithm. (B) Fingerprint analysis scheme. Different identifiability matrices were built in order to investigate the functional connectomes (FCs) identifiability in healthy subjects (HS) and patients with Parkinson’s disease (PD). Correlating test-retest individuals’ FCs we obtained the blue and the red boxes, that represents the identifiability characteristics of HS and PD, respectively. Cross correlating test and retest FCs of subjects of different groups we obtained hybrid identifiability matrices. From these matrices we were able to calculate the similarity of each patient’s FC with respect to the ones belonging to the healthy group (I*clinical* score).

### Connectome fingerprint

The difference between identifiability of both PD and HS at the level of the whole dataset was investigated, comparing the identifiability parameters of the two datasets. To check if significant differences were observed, we permuted 10000 times the labels of the participants (i.e., HS and PD), and each time we computed the difference in the parameters between the two groups (see methods for further details). Permutation testing was performed within each frequency band, but only the beta band showed significant differences, after applying False Discovery Rate (FDR) correction^16^ (Fig. 2). Specifically, HS displayed higher I-diff (HS = 0.33 ± 0.21; PD = 0.24 ± 0.22; pFDR = 0.034), I-self (HS = 0.42 ± 0.23; PD = 0.3 ± 0.23; pFDR = 0.017), and I-others (HS = 0.09 ± 0.04; PD = 0.6 ± 0.2; pFDR < 0.001) scores compared to PD patients. As a whole, patients showed lower differential identifiability (i.e., I-diff). Moreover, it is noteworthy that healthy individuals displayed high self-similarity despite being more similar among themselves with respect to patients.

**Figure 2.**
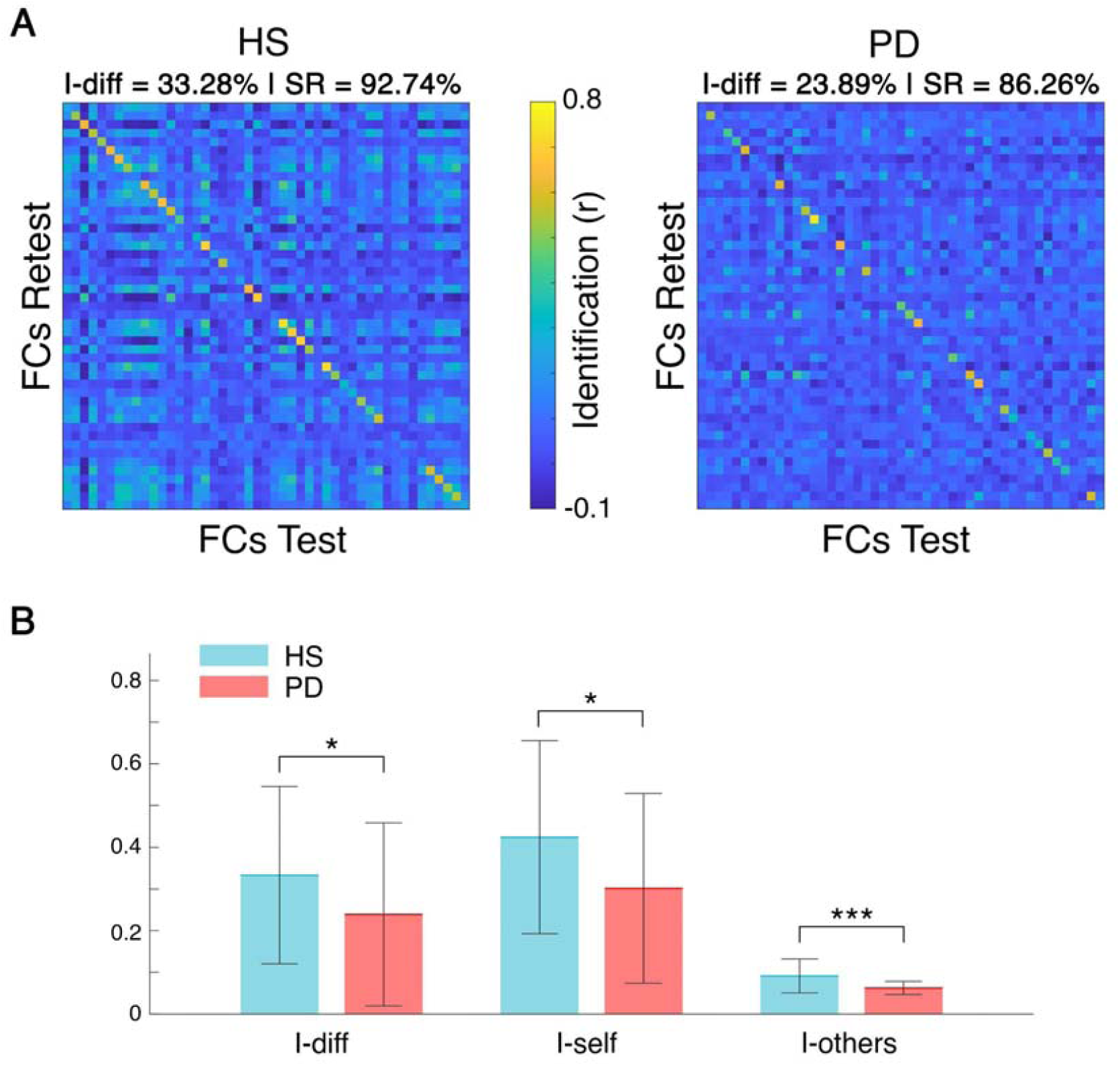
Brain identification in healthy and Parkinson’s disease. (A) Identifiability matrices of healthy subjects (HS) and patients with Parkinson’s disease (PD). The main diagonal is representative of the self-identifiability (I-self), while off-diagonal elements are representative of the similarity among different individuals (I-others). The difference between those values is described as differential identification (I-diff) and gives an estimation of the fingerprinting level of a group. These matrices are based on the functional connectomes computed in beta band. Note that the more the main diagonal is visible, the more the subjects turn out to be identifiable. Success Rate (SR) is reported too, as a percentage of the number of times an individual is recognizable with respect to other individuals within the same group. (B) Statistical comparison between fingerprint parameters calculated on the identifiability matrices of HS and PD. HS shows higher identifiability with respect to PD. Significance p-value: *p□<□0.05, **p□<□0.01, ***p□<□0.001.

### Edge-based identifiability

To assess the contribution of individual FC’s edges in determining the level of fingerprinting^13^ we exploited the intraclass correlation (ICC) statistics (see methods for further details). We observed two different behaviors in HS and PD groups (Fig. 3). Firstly, in healthy individuals many edges have high values of ICC, hence contributing to the identification, while in the patients group the edges have generally lower values, and a few, scattered edges contribute to the identifiability. All in all, this analysis indicates a more stable edges’ connectivity in healthy subjects, across the test-retest sessions.

**Figure 3.**
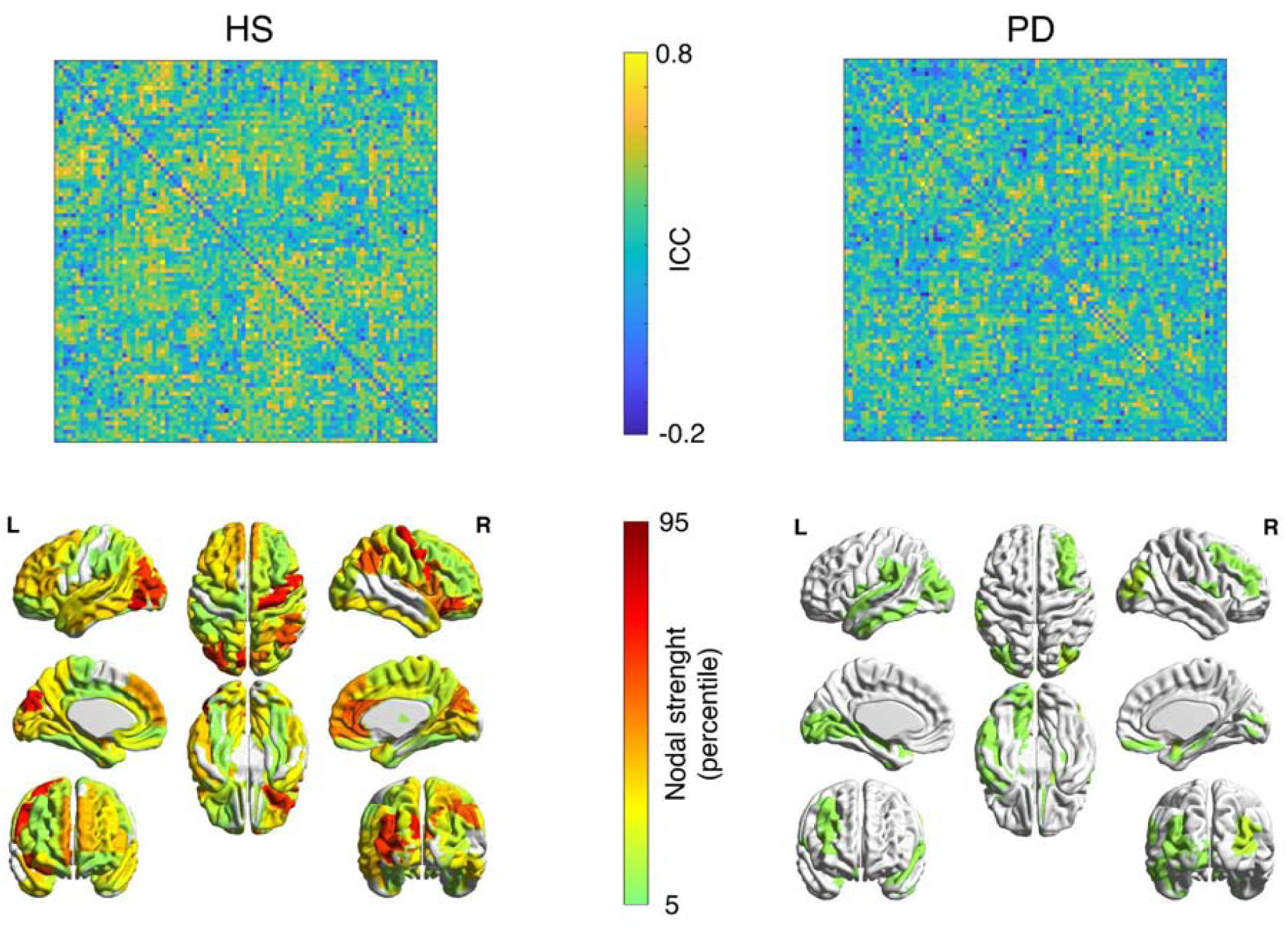
Edge contribution to connectome fingerprint. Intra-class correlation (ICC) for the beta band connectivity, assessing the brain regions contribution to identifiability. Higher ICC values of an edge means major contribution of that edge to the identifiability. The same results are shown as brain renders displaying the nodal strength of most reliable edges (above the 75 percentile of the distribution; colorbar borders represent the 5 and the 95 percentile).

Moreover, we calculated the average success rate (SR) value of each group, by investigating the percentage of identifiability of each subject within its own group. Hence, we analyzed the distribution of SR values in the fingerprint analysis performed adding 100 edges per iteration, from the most to the least stable ones, according to the ICC matrices of each group. Figure 4 shows that the HS group immediately reached a complete SR (100%), with a result that remained stable when adding more edges, and dropped starting from ∼2300 edges. Conversely, the PD group’s SRs did not reach the same values of the HS group, and the values never reached a stable level, slowly decreasing, until markedly dropping after ∼2800 edges. As a reliability test to our approach, we performed a surrogate analysis, this time adding edges in random order, obtaining a null distribution of SR values that were to be expected given a random selection of edges (Fig. 4, left panel). As evident, the SR was always above chance level, thereby showing that the selected edges carry relevant information to identify subjects. Furthermore, we observed the SR values distribution of the PD group, when ordering the regions according to the ICC matrix of the HS (Fig. 4, right panel). In this case, the patients’ SR values dropped compared to the distribution performed according to the ICC matrix of the patients themselves, and was nearly invariably within the null distribution, hence confirming that edge specificity is lost in PD.

**Figure 4.**
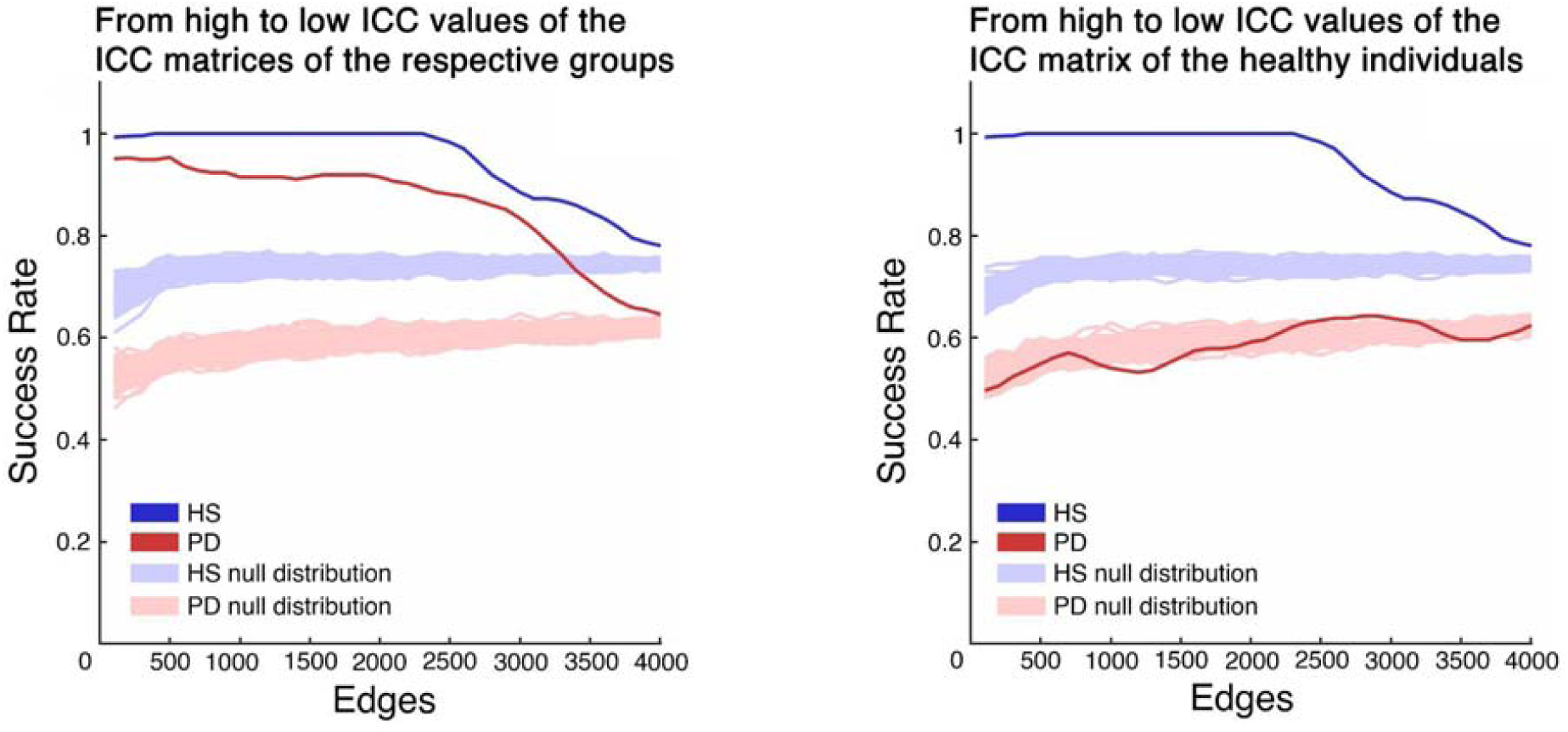
Identifiability based on the edge contribution. Success rate (SR) distribution in identifying individuals when performing fingerprint analysis including 100 edges at a time. SR distributions of healthy subjects (HS, blue line) and Parkinson’s disease patients (PD, red line), were obtained adding the edges from the most contributing to the least contributing to identifiability, relying on the ICC values. Actual distributions were compared to their respective null distribution (light blue for HS, and light red for PD) obtained repeating the same analysis one hundred times, including the edges in a random order. The left panel shows the analysis performed using the ICC matrices belonging to each group. The right panel shows the analysis performed considering the ICC matrix of the healthy individuals for both HS and PD group.

### Clinical features of fingerprinting

Finally, in order to verify the clinical value of this approach, we tried to predict the UPDRS scores, relying on the I*clinical* values in the beta band. To this end, we built edge-based multilinear models including the I*clinical* calculated with a growing subset of edges (from 100 to full FC, adding 100 more at each iteration) as predictor. Age, education, gender and disease duration were added as predictors as well, while the UPDRS was set as response variable. The highest similarity between actual and predicted UPDRS scores was observed at 500 edges (Spearman ρ = 0.59). Indeed, the model explained 44% of the variance of the UPDRS (R2 = 0.44) (Fig. 5 left panel). Both the I*clinical* (p = 0.0005, β = - 144.72) and the disease duration (p = 0.001, β = 0.14) significantly contributed to the predictions. Individual predictions and the distribution of the residuals obtained through the k-fold validation method^17^ are shown in Figure 5 (mid and right panels respectively). No significant contribution of age, education, and gender was observed.

**Figure 5.**
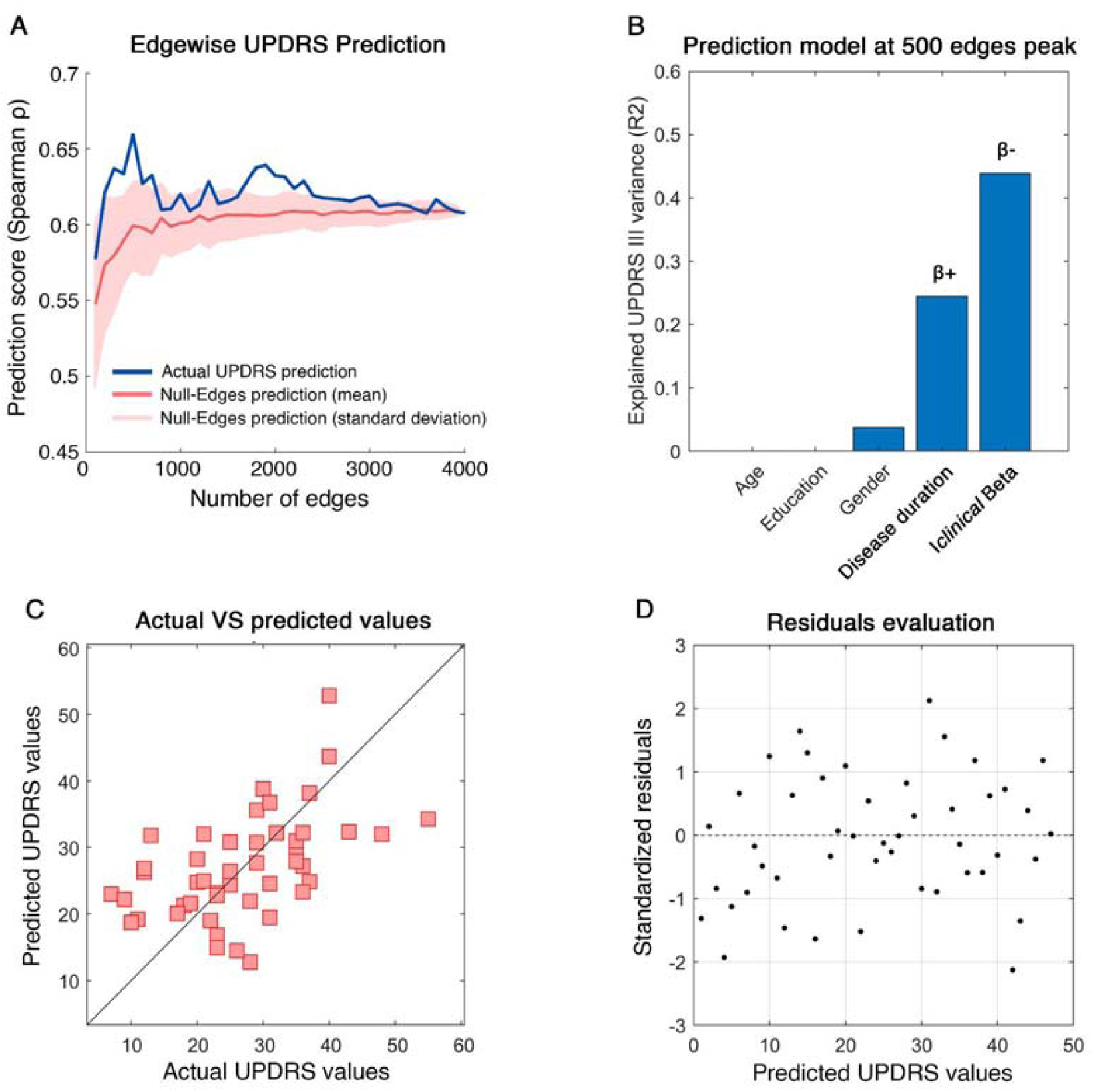
Motor impairment prediction based on “clinical fingerprint”. The analysis aims to predict the motor impairment of the patients assessed through UPDRS, relying on the clinical identifiability (I*clinical*) score. **A)** Edges are added iteratively (100 per time up to whole-brain) based on the Parkinson’s disease (PD) patients’ ICC values, from the most to the least contributing to identifiability (x axis). Hence, the prediction performance (k-fold cross validation with k = 5) of each multilinear model based on the I*clinical* is evaluated as the Spearman correlation coefficient (Spearman’s ρ, on y axis) between actual and predicted UPDRS values (blue line). For comparison, we built a null model obtained by repeating the same analysis 100 times, but selecting the edges randomly (the red line represents the mean prediction of the null model; the shaded red area represents the standard deviation of the null model predictions). The following panels show the results of multilinear model with the highest performance, i.e., when the I*clinical* is calculated considering the 500 most reliable edges for PD patients’ identification (ICC score). **B)** The panel shows the variance explained by the additive model including five variables (age, education, gender, disease duration, and I*clinical* in beta band). Significant predictors in bold; positive/negative coefficients indicated with β+/β-. **C)** The panel shows the correspondence between the actual UPDRS values and the ones predicted by the model. **D)** The panel displays the distribution of the standardized residuals.

Furthermore, we specifically observed the relationship between the clinical fingerprint in the beta band and the motor condition of our patients. The correlation test between the I*clinical* at 500 edges peak and the UPDRS scores highlighted a significant negative correlation between the two parameters (r = -0.48, p < 0.001), as shown in Figure 6.

**Figure 6.**
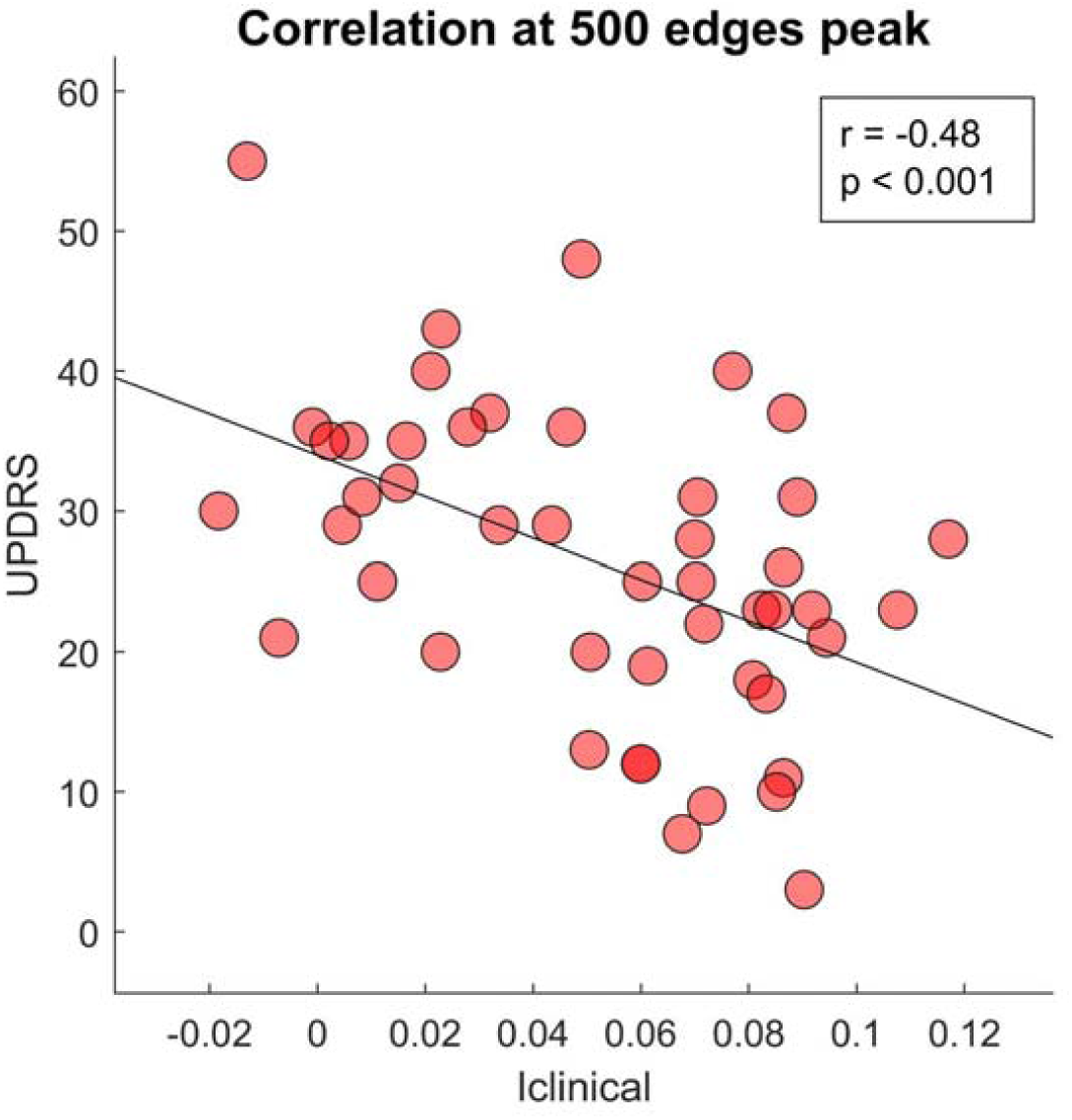
Relationship between motor impairment and clinical identifiability. Pearson’s correlation between motor impairment assessed through UPDRS and clinical identifiability expressed by I*clinical* score in beta band. The Iclinical was computed including the 500 most reliable edges according to the ICC scores. The negative significant coefficient indicates better motor condition (low UPDRS values) when patients connectivity is more similar to the healthy individuals (high I*clinical*), and vice versa.

## DISCUSSION

In this study, we set out to investigate whether the changes induced by Parkinson’s disease in the large-scale brain connectivity could reflect into reduced connectome-based identifiability of patients. We tested this hypothesis within the recently developed framework of the clinical connectome fingerprinting^18^, comparing the identifiability of patients with PD and matched controls. We therefore extracted a clinical fingerprinting score for each patient^13^, and exploited it to predict motor impairment in each patient, working under the hypothesis that lower identifiability would be linked to dysregulated functional connectivity.

The fingerprinting analysis was conducted by comparing the connectomes in the test/retest sessions of the participants, in each group (i.e., PD and HS) separately. The connectomes were calculated using the PLM to measure synchronization^14^ in each of the canonic frequency bands. Significant results were exclusively found in the beta band. As mentioned above, the beta band is consistently reported as altered in PD^5^. Our results showed that the PD patients displayed lower differential identifiability compared to the healthy individuals (Fig. 2). Hence, the healthy connectomes are more recognizable and subject-specific. In detail, the patients showed both lower I-self and I-others scores. On the one hand, the lower I-self indicates lower similarity between the two connectomes belonging to one subject. On the other hand, the reduced I-others represents greater heterogeneity among the connectomes of the PD group. However, the I-self showed greater reduction as compared to the I-others. We speculated that this feature might be caused by the loss of the subject-specific fine-tuning of large-scale dynamics, which reflects itself primarily on the loss of similarity of a subject with him/herself, rather than of a subject with the other subjects. In PD, dopamine depletion is able to alter the brain network organisation and its dynamics, and this approach may be able to catch one of the features of the unstable brain activity occurring in patients’ connectomes^4,19–21^. Indeed, the reduced similarity of subsequent MEG recordings may be a consequence of the dysregulated brain activity occurring in the beta band in PD^7^.

Furthermore, the edge-specific reliability of the connectomes dropped drastically in PD patients, and reflected itself in much lower nodal stability (Fig. 3). It is crucial to note that ICC matrices show the reliability of the communication between brain regions, but not the magnitude of the connectivity itself. Hence, we retrieve an information that is complementary to the one related to connectivity. In summary, our results suggest that several brain regions contribute to the identifiability in the healthy controls, while only a few do so in PD. In particular, it can be observed that the motor regions provide a major contribution to the identifiability of healthy individuals, but not to that of PD patients. Once again, the abnormal activity patterns clustered in the motor regions, which are particularly affected in PD patients (Fig. 3), may lead to the reduced contribution to the identifiability. To date, the quality of the alterations (in the sense of hypo/hyperactivation) related to the regions involved in the PD motor patterns is still debated^22^. With regard to the motor cortex, several studies reported controversial results when comparing PD patients and healthy controls. Indeed, using functional MRI, Buhman et al.^23^, and Grafton^24^ showed hypoactivation, while Haslinger et al.^25^, and Sabatini et al.^26^ reported hyperactivation. However, our approach did not focus on the connectivity itself, rather we estimated how stable it is across multiple recordings of subjects belonging to the same group. Nevertheless, even in this case beta band activity in PD connectomes revealed alterations, in terms of stability and identifiability.

Then, we observed the behavior of identifiability as a function of the number of edges utilized to perform it. In both groups the identifiability was higher when only taking into account a subset of edges, and not the complete functional connectome. Furthermore, following the inclusion of a minimum number of edges, the healthy individuals reached a complete and stable subject recognition, while the patients lacked to do so. A possible explanation would be that the full connectome contains more redundant information, i.e., patterns that are not subject-specific but, rather, shared by multiple subjects (Fig. 4). Indeed, several studies focused on the analysis of patterns of inter-subject variability, reporting the presence of a global common organization in conjunction with subject-specific patterns^27–29^. Hence, considering that there is a concordance across subjects over the edges that contribute to the identification, one might further speculate that subject-specific information is contained preferentially in specific functional patterns. Furthermore, the altered activity in the beta band might contribute to the lack of identifiability in the PD group^30^. In other words, impairment of the fine-tuned regulation of the large-scale activity of the brain might make the system unable to keep its (presumably) optimal trajectory. This would in turn result in higher variability and, thus, reduced individual identifiability. Indeed, this result is in accordance with our previous interpretation of the ICC matrices (Fig. 3), where we note that in healthy individuals there is a broad contribution to identifiability from several brain regions, in contrast to the PD group which shows fewer stable edges. This would even provide an interpretation as to why the identifiability in the healthy subject remains stable when a large number of connections is added into the analysis.

Subsequently, we wondered if the altered fingerprint of PD subjects could be related to the clinical picture of the disease. Similarly to a previous study^13^, we used the I*clinical* score in beta band to predict the motor impairment typical of PD, as assessed using the UPDRS. In particular, we observed that 500 edges was the number of edges that maximized the prediction. We found that our model could explain nearly 44% of the variance of the UPDRS scores across individuals. Even when accounting for nuisance variables such as age, level of instruction and disease duration, the I*clinical* significantly improved the performance of the model. Besides the I*clinical*, disease duration also contributed to the predictions. Given the negative beta-coefficient of the I*clinical*, we can conclude that the more the identifiability of a PD patient’s connectome is similar to that of the HS group, the milder its motor impairment. Noteworthy, these results were validated with a k-fold cross-validation that reduced the risk of overfitting.

It should be noticed that the edges that were relevant for the clinical identification were also the ones responsible for the prediction of the UPDRS. In both edge-based analyses (i.e., identification and clinical motor score prediction), the edges were ordered in the same way and in both cases the best performance was obtained considering only a few hundreds of edges. Hence, there is substantial overlap between the edges that allow identification and those that allow the clinical prediction. This points toward the clinical validity of this approach, showing that the selected edges were related to a functional outcome (i.e., the UPDRS score). This result once more supports the idea that the loss of stability that leads to lower identifiability might be related to mechanisms that are pathophysiologically relevant in PD. In fact, several studies showed a correlation between brain connectivity features in the beta band and motor impairment^31,32^. Furthermore, we also demonstrated the inverse linear relationship occurring between the I*clinical* in the beta band and the UPDRS scores. Our results are in line with these findings and demonstrate that in PD, the connectome-based identifiability conveys the severity of the disease. Since the UPDRS is one of the most reliable and used motor scales in the clinical settings^33–35^, we believe that a scalar score, based on the whole connectome conveying subject-specific features of the brain functional connectivity may be of help in the management of the disease.

In conclusion, we applied the fingerprint approach to PD, showing that the subject-specific brain network recognition is linked to the clinical condition. Firstly, we highlighted the lower identifiability of the patients with respect to the healthy individuals. Furthermore, we showed that the degree of the connectome-based identifiability of the patients (with respect to the healthy population) is related to their clinical motor condition. Importantly, all the results were observed within the beta band, which is known to be highly involved in PD. We believe that these results will contribute to the optimization of the diagnosis and treatment of PD, allowing the inclusion of information about the patient-specific dysregulation of brain activity on the large-scale.

## METHODS

### Participants

We recruited forty-seven patients (30 males, 17 females) affected by PD, with a mean age of 65 years (±9.7), and a mean education of 11.3 years (±4.2). The diagnosis of PD was fulfilled in accordance with the United Kingdom Parkinson’s Disease Brain Bank criteria^36^. Inclusion criteria were: (a) PD onset after the age of 40 years, to exclude early onset parkinsonism; (b) a modified Hoehn and Yahr (H&Y) stage□≤□2.5. Exclusion criteria were: (a) dementia associated with PD according to consensus criteria; (b) any other neurological disorder or clinically relevant medical condition. Disease severity was assessed through a motor examination in “off-state”, using both the UPDRS^15^, and the Hoehn and Yahr (H&Y) stages^37^. Forty-seven healthy subjects (HS) were recruited as well, matched for gender (30 males, 17 females), age (61.8 ±10 years), and education (12.9 ±4.6 years). All the recruited individuals were right-handed. The study was performed in accordance with the Declaration of Helsinki, and all the participants signed an informed consent. The local Ethic Committee of University of Naples “L. Vanvitelli” approved the study.

### MEG acquisition

All the participants underwent a MEG scan. The system was composed of 154-magnetometers, and 9 reference sensors. The scan is placed in a magnetically shielded room (AtB Biomag UG, Ulm, Germany). Before each acquisition we used Fastrak (Polhemus) to record the position of four position coils and four reference points, located on the head of the participant (nasion, right and left pre-auricular, and apex)^38^. This allowed us to locate the position of the head during the acquisition, activating the coils before each segment of registration. Participants were recorded during resting state with eyes closed. Two recordings, 3.5 minutes long, were separated by a short break roughly 2 minutes long. This amount of time is a tradeoff which allows to record enough signal and prevent drowsiness of the subject^39,40^. Electrocardiographic (ECG) and electro-oculographic (EOG) signals were acquired in order to remove physiological artefact^40^. Signals were sampled at 1024 Hz after anti-aliasing filtering.

### Magnetic resonance imaging

Eighty participants underwent magnetic resonance imaging (MRI). A 1.5-T Signa Explorer scanner equipped with an eight-channel parallel head coil (GE Healthcare, Milwaukee, WI, USA) was used. Specifically, three-dimensional T1-weighted images (gradient-echo sequence Inversion Recovery prepared Fast Spoiled Gradient Recalled-echo, time repetition = 8.216 ms, TI = 450 ms, TE = 3.08 ms, flip angle = 12, voxel size = 1 × 1 × 1.2 mm3; matrix = 256 × 256) were recorded. From the total of 94 subjects recruited, 7 patients and 7 healthy individuals refused/were unable to undergo MRI, and a standard template was used for source reconstruction.

### Preprocessing

Preprocessing and source-reconstruction was performed similarly as in Sorrentino et al.^41^. In short, a 4^th^-order Butterworth IIR band-pass filter was implemented, using the Fieldtrip toolbox in MATLAB^42^, in order to filter the data in the 0.5-48 Hz range. Then, through the Principal Component Analysis^43^, we orthogonalized the signals with respect to the reference signals. Then, an experienced rater identified and removed noisy signals and segments after visual inspection. Finally, supervised Independent Component Analysis^44^ was performed to identify the ECG and, if present, the EOG components present in the MEG signals.

### Source reconstruction

We co-registered the MEG data with the native MRI of each subject. Then, we obtained the time series of 116 regions of interest (ROIs), based on the AAL atlas^45^, using the volume conduction model proposed by Nolte^46^, and applying the Linearly Constrained Minimum Variance^47^ beamformer algorithm included in the Fieldtrip toolbox^42^. The resulting time series were band-pass filtered in each canonical frequency band (i.e., delta (0.5 – 4 Hz), theta (4 – 8 Hz), alpha (8 – 13 Hz), beta (13 – 30 Hz), and gamma (30 – 48 Hz)). Only 90 ROIs were selected for further analysis, since we excluded ROIs related to the cerebellum because of low reliability.

### Synchrony estimation

Synchronization was estimated through the PLM, measuring the phase difference in time between brain regions^48^. In short, the PLM is based on the spectrum of the interferometric signal between pairs of brain regions, and it is unaffected by volume conduction. Its values range from 0 (no synchronization) to 1 (synchronization). Computing the PLM between each couple of regions we obtained, per each frequency-band, two functional connectomes (one per recording segment), that we named test and retest.

### Fingerprint analysis

To evaluate the fingerprinting in our population, we employed an approach based on functional connectomes, as originally proposed in Sorrentino et al.^13^. Firstly, we aimed to create an Identifiability matrix^18^ (Fig. 1). The IM features the participants on rows and columns, while the entries are the Pearson’s correlation coefficient between the test and retest FCs of each participant. The IM contains information on homo-similarity (or self-similarity – I-self, the main diagonal elements) and hetero-similarity (or similarity of each subject with the others – I-others, off diagonal elements). The I-self conveys the similarity between the connectomes derived from the test and re-test sessions of the same participant. The I-other conveys the similarity between the test session of one participant with the re-test sessions of the other participants. Computing the difference between the mean I-self and the mean I-others, we can obtain the differential Identifiability (I-diff)^13,18^. This score offers an estimation of the fingerprint level of a specific dataset. Furthermore, we calculated the success rate value, to determine the percentage of identifiability of the subjects within a group. It was computed observing the percentage of times that each subject displayed an I-self value higher than the I-others values tested on the corresponding row and column. Finally, crossing the test-retest FCs of the healthy individuals and of the patients, it is possible to obtain the I*clinical* score (“clinical identifiability”, or “clinical fingerprint”). To this end, two hybrid IMs are built. The first matrix is obtained correlating the healthy test FCs and the clinical retest FCs, while the second matrix is computed correlating the healthy retest FCs and the clinical test FCs. From each matrix we can calculate, for each patient, the average similarity with the controls. Averaging the score obtained from the two hybrid matrices, we obtain an I*clinical* score for each patient^13^. This is an estimate of how harder it is to identify patients as compared to healthy controls. For further details, please refer to Sorrentino et al.^13^.

### Region of interest for fingerprint

Borrowing from previous work on identifiability^18^, we used the intraclass correlation coefficient (ICC)^49^ to quantify the edgewise reliability of individual connectomes. Basically, the ICC quantifies how much the elements belonging to the same group are similar to each other (higher ICC values correspond to higher similarity). In our case, edges display high ICC values if they keep similar values of synchronization across the test-retest sessions. Hence, edges that are steady might contribute more to the overall identifiability. Then, we performed the fingerprint analysis sequentially adding edges according to their ICC values. We added 100 edges at each iteration, and computed the SR values at each iteration. To check that the selected edges were in fact the most relevant ones in terms of identification, we built a null model for each group (HS and PD), selecting 100 times at each iteration a set of randomly chosen edges, and then performing the fingerprint analysis on those. This way, we build a null distribution of the SR values to be expected by selecting edges randomly.

### Edges of interest for clinical predictions

Furthermore, we tested the hypothesis that the I*clinical* score based on a subset of edges could predict the clinical condition of the patients. Hence, we built a multilinear regression model to predict the UPDRS scores from the I*clinical* values^50^. We added more covariates to the model, in order to account for the effect of age, education, gender, and disease duration. Multicollinearity was assessed through variance inflation factor (VIF)^51,52^. We strengthened the reliability of our model using the k-fold cross-validation with k=5 ^17^. Specifically, k iterations were performed and at each iteration the k^th^ subgroup was used as a test set. To check if a subset of edges were mostly responsible for the prediction of the UPDRS we adopted a similar approach as before, and computed the same model multiple times. At each iteration, 100 edges were added according to their ICC value (in descending order), and the I*clinical* was computed. Furthermore, at each iteration, 100 surrogate models were computed each based on the I*clinical* computed on a random selection of edges. At each iteration the Spearman’s correlation coefficient between predicted and actual UPDRS values was calculated and considered as a prediction score.

### Statistics

Statistical analysis was carried out in MATLAB 2020a. I-self, I-others and I-diff values were compared between the two groups. The comparisons were performed through permutation testing, by randomly rearranging the labels of the two groups 10,000 times^53^. The absolute value of the difference was computed at each iteration, obtaining a distribution of the randomly determined differences^54^. This distribution was compared to the observed differences to retrieve a statistical significance. The possible relationships between variables were investigated using Pearson’s correlation. Results were corrected by False Discovery Rate (FDR) correction^16^. Significance level was set at p-value < 0.05 after correction.

## Data Availability

All data produced in the present study are available upon reasonable request to the authors

## Acknowledgments

G.S. acknowledges financial support from University of Naples “Parthenope” within the Project “Bando Ricerca Competitiva 2017” (D.R. 289/2017) and the Project “Ricerca Locale”, 2018. V.J. and P.S. acknowledges financial support from European Union’s Horizon 2020 research and innovation program under grant agreement 945539 (SGA3) Human Brain Project and from grant agreement No. 826421 Virtual Brain Cloud.

## Author Contributions

T.L.E. conceived the idea, wrote the codes in Matlab, including those for the null model, the regression and the statistical analysis, and wrote the original draft of the manuscript. M.R. deigned the study, created the figures, performed the analyses, and wrote the original draft of the manuscript. L.M. and P.A. performed the MEG recordings, cleaned the signals and processed the MEG data (beamforming and source reconstruction). R.A. performed the statistical analyses and created the figures. D.M.R. and T.A. enrolled and screened the patients. L.F. provided valuable feedback for the manuscript. A.E. wrote the code for the fingerprint analysis and provided valuable feedback for the manuscript. S.G. provided the funding, enrolled, screened and performed MRI of both patients and healthy controls, provided valuable feedback for the manuscript. J.V. provided the funding and provided valuable feedback for the manuscript. S.P. enrolled and screened the patients and the healthy controls, supervised all the pipeline (recording, cleaning, beamforming, source reconstruction, data analysis), wrote and revised the original draft of the manuscript. All authors have read and approved the final manuscript.

## Data and materials availability

All data needed to evaluate the conclusions in the paper are present in the paper and/or the Supplementary Materials. Additional data can be provided by the corresponding author, upon reasonable request.

